# Change in Social Participation of Adults with Spinal Cord Injury During the First Two Waves of the COVID-19 Pandemic in Canada

**DOI:** 10.1101/2024.09.30.24314551

**Authors:** Noémie Fortin-Bédard, Félix Nindorera, Jean Leblond, Caroline Rahn, Krista L. Best, Jaimie Borisoff, Shane N. Sweet, Kelly P. Arbour-Nicitopoulos, François Routhier

**Affiliations:** Centre for Interdisciplinary Research in Rehabilitation and Social Integration, Centre intégré universitaire de santé et de services sociaux de la Capitale-Nationale, Quebec City, Canada; School of Rehabilitation Sciences, Université Laval, Quebec City, Canada; British Columbia Institute of Technology, Vancouver, Canada; Department of Kinesiology and Physical Education, McGill University, Montréal, Canada; Center for Interdisciplinary Research in Rehabilitation of Greater Montreal, Montreal, Canada; Faculty of Kinesiology and Physical Education, University of Toronto, Ontario, Canada

**Keywords:** COVID-19, Spinal cord injury, Manual Wheelchair, Social Participation, Rehabilitation, Social Support, Pandemic

## Abstract

**Introduction:** The change in environmental and social context measures during the COVID-19 pandemic affected daily activities of people with spinal cord injury (SCI), their interactions within the community, and consequently their social participation during the first wave of the pandemic. However, there is little information about the changes in social participation as the pandemic evolved in Canada. Objective: To describe the change in social participation of adults with SCI after the first two years of the COVID-19 pandemic in Canada.

**Methods:** A follow-up from a previous study exploring the social participation of adults with SCI living during the first wave was conducted eight months later (second wave). Social participation was measured using the Assessment of Life Habits (LIFE-H 4.0) and Measure of Quality of the Environment (MQE) among 18 adults with SCI.

**Results:** Participants reported increases between both waves of COVID-19 in life habits categories, including mobility, personal care and health, nutrition, and recreation. New environmental factors were identified as facilitators, including the increased availability of businesses in the community.

**Conclusion:** These findings suggest that people with SCI increased fulfillment and satisfaction of certain life habits. Reduced environmental barriers and increased facilitators improved social participation as the pandemic evolved.

## 1. Introduction

In 2022, 8 million Canadians of 15 years and older were living with one or more disabilities (Statistics Canada, 2022). According to the World Health Organization, the COVID-19 pandemic and the associated protective and isolation measures have highlighted significant inequities within the population, including those experienced by people with disabilities (PWD) (World Health Organization, 2022). In this state of emergency context, the rapid changes (e.g., lockdown, changes in the supply of health services, and closure of some businesses) (Canadian Institute for Health Information, 2022) had an impact on physical activity levels, precarious employment and deteriorating mental health for PWD (Brown & Ciciurkaite, 2023; Lebrasseur et al., 2021; Tuakli-Wosornu et al., 2023). This finding is particularly concerning considering that PWD often have poorer health outcomes than people without disability due to the multiple barriers that limited their accessibility to healthcare services, even before the pandemic (World Health Organization, 2022). These barriers include, for example, physical environment, logistic related to transportation to the healthcare centers and lack of understanding of their healthcare needs (Hashemi et al., 2022). Previous studies have reported that the pandemic increase the social isolation experienced by PWD (Kersey, 2022). Thus, the pandemic has inevitably affected the way PWD carry out their daily activities and how they interact with their community (Fortin-Bédard et al., 2022). In turn, PWD experienced changes to their social participation (Levasseur et al., 2022), and an overall reduction in their quality of life (Friedman, 2021).

According to the World health organization, the annual incidence of spinal cord injury (SCI) is estimated around 40 and 80 cases per million inhabitants worldwide (World Health Organization, 2013). People with SCI represent a significant group within the range of PWD, with nearly 31 000 individuals living with an SCI in Canada in 2019 (Thorogood et al., 2023). SCI is a major cause of long-term disability, accounting for over 4.5 million years of life lived with disability worldwide in 2021 (Benabid et al., 2019; Ding et al., 2022). Findings from a systematic review showed a wide range of the consequences of SCI (Post et al., 2010), such as loss of motor functions and bowel dysfunction (World Health Organization, 2013). According to the *Human Development Model - Disability Creation Process* (HDM-DCP), due to interaction between characteristics of the individuals and the environment (i.e., obstacles or facilitators), people with SCI are susceptible to face disabling situations (International Network on the Disability Creation Process). Therefore, the presence environmental facilitators are important to ensure the social participation for people with SCI (Post et al., 2010). For example, people with SCI have specific need to maintain their social participation, such as peer support, adapted transportation, technical aids and healthcare SCI-related services (Beauregard et al., 2012). As the pandemic progressed in Canada (Canadian Institute for Health Information, 2022), health and protective measures were added, modified, and in some cases, removed, leading to a period of uncertainty and adaptation for PWD, such as people with SCI (Fortin-Bédard N et al., 2023). In a study conducted among 382 adults with SCI, 59% of participants reported that the COVID-19-related restrictions negatively affected their social participation (Krause & Jarnecke, 2023). More specifically, 51% agreed or strongly agreed that the COVID-19 restrictions reduced drastically their community participation (Krause & Jarnecke, 2023). In addition, a recent scoping review (2024) revealed that the pandemic was detrimental to healthcare utilization, access, and outcomes in individuals with SCI (Senthinathan et al., 2024). For example, the frequency of going out, receiving home-visit nursing, and rehabilitation services has decreased during the pandemic period (Matsuoka & Sumida, 2022). In Canada a study reported that protection measures (e.g., foot pump for hand sanitizers) for people with spinal cord injury (SCI) were not adequately adapted during the first wave of the pandemic (Fortin-Bédard et al., 2022). However, positive attitudes, support from family and friends, and availability of businesses in the community (e.g., grocery, drug and hardware stores) were reported as facilitators to social participation for people with SCI at the beginning of the pandemic (Fortin-Bédard et al., 2022).

Since social participation has been positively associated with personal fulfillment and a sense of wellbeing and belongingness (Best et al., 2022; Couture et al., 2020). These findings are important. Indeed, even before the pandemic, PWD faced obstacles to attaining full social participation, such as discriminatory attitudes (Hästbacka et al., 2016), problems with transportation (Lidal et al., 2007), lack of facilities, and difficulties with accessibility (Jaarsma et al., 2014). Furthermore, social participation may also have evolved for people with SCI since the first wave of the pandemic. The changes in social participation throughout the pandemic are less well documented than social participation at specific times. In this context, the objective of this study was to describe the change in social participation of adults with SCI after the first two years of the COVID-19 pandemic in Canada. We hypothesized that facilitators to social participation identified earlier in the pandemic may no longer be present and that new barriers may have ensued as the pandemic continued.

## 2. Materials and Methods

### 2.1 Design and participants

This is a descriptive cross-sectional study representing a follow-up of a previous study exploring the social participation of adults with SCI living in Canada during the first wave (first wave, abbreviated T1 hereafter) (Fortin-Bédard et al., 2022). The present study was conducted approximately eight months later (second wave, abbreviated T2 hereafter). This project was approved by local Research Ethics Boards (MP-13-2018-415). The standards for reporting observational studies (STROBE) were followed (von Elm et al., 2007). Participants were recruited from a convenience sample of an ongoing Canadian multi-site trial (i.e., Quebec City, Montreal, and Vancouver). All methodological details were previously described (Fortin-Bédard et al., 2022).

### 2.2 Data Collection

The sociodemographic data (i.e., age, sex, province of residence, employment status, length of time using a wheelchair) were collected at the baseline of the larger ongoing trial. In the present study (T2), each participant completed two validated questionnaires to measure social participation between December 2020 and April 2021 namely Assessment of Life Habits (LIFE-H 4.0) and Measure of Quality of the Environment (MQE). Of note, participants completed the same items that were selected from these two questionnaires during T1 (first wave) to measure changes in social participation (Fortin-Bédard et al., 2022).

### 2.3 Assessment of Life Habits (LIFE-H 4.0)

The LIFE-H 4.0 was used to measure the quantity and the quality of social participation according to the conceptual HDM-DCP (Fougeyrollas P. & Noreau L., 2014). We selected thirty-two items from the LIFE-H 4.0 to assess the realization of twelve life habits. In the LIFE-H, these twelve life habits are divided into two broad categories: day-to-day habits and social roles habits. The two categories are further divided into six life habits each: 1) communication; 2) mobility; 3) nutrition; 4) physical fitness and psychological well-being; 5) personal care and health; 6) housing; 7) responsibility; 8) interpersonal relationships; 9) community and spiritual life; 10) education; 11) employment and, 12) recreation (Fougeyrollas P. & Noreau L., 2014) (Supplemental Table 1). For each item, scoring is applied according to a series of questions, including the realization of the life habits. A total score of the level of social participation was calculated based on the answer to the 32 items. Total scores were then rescaled over ten points for comparison purposes. A higher score indicates greater independence, less difficulty, and greater satisfaction. Of note, all participants reported that the education life habit category of the LIFE-H was not applicable. Thus, the education life habit category is not shown.

**Table 1.** Description of sociodemographic information and participant characteristics^1^.

| Characteristics | n (%) |
| --- | --- |
| Age, years (mean [SD]) | 48.8 [15.1] |
| Sex |  |
| Women | 6 (33.3) |
| Men | 12 (66.7) |
| Province of Origin |  |
| Quebec | 14 (77.8) |
| British Columbia | 4 (22.2) |
| Employment Status<sup>1</sup> |  |
| Employed | 3 (16.7) |
| Unemployed | 9 (50.0) |
| Retired | 5 (27.8) |
| Highest Education Level |  |
| Less than high school | 0 (0.0) |
| High school (started) | 6 (33.3) |
| College/University (started) | 2 (11.1) |
| College/University (completed) | 7 (38.9) |
| Post-graduate studies | 2 (11.1) |
| Annual Household Income, \$1 | |
| < 14'999\$ | 0 (0.0) |
| 15'000-29'999 | 7 (38.9) |
| 30'000-44'999 | 3 (16.7) |
| 45'000-59'999 | 3 (16.7) |
| 60'000-74'999 | 1 (5.5) |
| 75'000 > | 2 (11.1) |
| Diagnostic |  |
| Paraplegic | 15 (83.3) |
| Tetraplegic | 3 (16.7) |
| Time Using Any wheelchair, years (mean [SD]) | 15.1 [13.3] |

<sup>1</sup>Missing information for one participant

### 2.4 Measure of Quality of the Environment (MQE)

The MQE was used to assess environmental barriers and facilitators to social participation (Fougeyrollas P. et al.). The tool was developed on the disability creation model and was content validated by a group of rehabilitation professionals. Participants self-responded to 45 items that were pre-selected by expert researchers to be indicative of a situation or factor that could influence daily life. Participants rated whether it was a facilitator, or an obstacle based on a 7-point Likert scale ranging from –3 (major obstacle); to 3 (major facilitator). Responses options, “I don’t know” or “Does not apply” were also possible. Reliability test (test-retest) was conducted among young adults with cerebral palsy, indicating that 85% of the items obtained agreement above 60% (Boschen, 1998).

### 2.5 Data Analysis

Descriptive analysis were conducted for sociodemographic data. Given that LIFE-H and MQE are ordinal scales, data were analyzed using a non-parametric rank-based repeated-measures ANOVA (package nparLD 2.1, R software 3.3) (Noguchi et al., 2012). Technically, this nparLD ANOVA is an extension of the Wilcoxon signed-rank test. It has the advantage of working with missing data (no imputation required, no exclusion of participants). The nparLD ANOVA also generates an effect size measure named Relative Treatment Effect (RTE), an adjusted probability. A RTE above (or below) .5 reflects that scores measured at a specific time are most often higher (or lower) than the median score of the whole dataset. To interpret effect sizes, the general boundaries by Vargha and Delaney (2000, see statistic A12 in Table 1 at page 106) were used for large (RTE≥.71 or RTE≤.29), medium (RTE≥.64 or RTE≤.36) and small (RTE≥.56 or RTE≤.44) effects (Vargha & Delaney, 2000). The nparLD ANOVA is appropriate for clinical data, since participants are often very similar at the initial time of measurement (e.g., a low realization of life habits for all) with an inflated variance at a second time since participants (e.g., unequally improved their capacities during the delay between measurements). Due to its non-parametric nature, the nparLD ANOVA has no assumption about sphericity of variance nor the stability of a distribution. The p-value significance was assumed at 0.05.

## 3. Results

Among the 14 participants who participated in the T1 study (Fortin-Bédard et al., 2022), 13 agreed to participate in T2. Five new participants who were not enrolled in the study at T1, participated in this study for a total of 18. Participants were predominantly male n=12 (67%), 48.8±15.1 years of age, and had 15.1±13.3 years of wheelchair (WC) experience (Table 1). Half of the participants were unemployed (50%) and lived in the province of Quebec (77.8%).

### 3.1 LIFE-H 4.0

Table 2 presents the change of all the life habits categories evaluated by the LIFE-H questionnaire. Between T1 and T2, the social participation score increased for the two broad categories of daily activities (p=0.0652; small effect size, 0.57) and social roles (p=0.0086; small effect size, 0.59; Table 2). More precisely, the life habits scores increased for categories of mobility (p=0.0285); personal care and health (p=0.0123); nutrition (p=0.0206); and recreation (p=0.0011). All effect sizes were rated small according to Vargha & Delaney (Table 2) (Vargha & Delaney, 2000). There was no statistically significant difference between T1 and T2 for the life habits category responsibilities, associative and spiritual life, housing, physical fitness and physiological well-being and health, employment, the interpersonal relationships, and communication (Table 2). Supplemental Figure 1 presents individual scores of the LIFE-H at T2 and Supplemental Figure 2 presents individual score changes of the LIFE-H between T2 and T1.

**Table 2.** Evolution of the life habits categories measured by the Assessment of Life Habits (LIFE-H) (Boucher et al., 2010)1.

| Life habits categories | Median score T1 | Median score T2 | p-value | Effect size (RTE)<sup>4</sup> |
| --- | --- | --- | --- | --- |
| Mobility | 5.3 | 6.4 | 0.0285 | 0.60 |
| Associative and spiritual life | 0.0 | 10.0 | 0.3303 | 0.56 |
| Housing | 7.6 | 8.0 | 0.6734 | 0.48 |
| Nutrition | 8.7 | 9.2 | 0.0206 | 0.58 |
| Physical fitness and physiological well-being | 7.0 | 8.8 | 0.1301 | 0.57 |
| Employment | 4.5 | 9.0 | 0.5854 | 0.52 |
| Recreation | 3.3 | 10.0 | 0.0011 | 0.60 |
| Communication | 10.0 | 10.0 | 0.4926 | 0.52 |
| Responsibilities | 9.3 | 9.3 | 0.7574 | 0.53 |
| Personal care and health | 6.9 | 8.0 | 0.0123 | 0.59 |
| Interpersonal relationships | 10.0 | 10.0 | 0.4996 | 0.48 |
| Day-to-day activities<sup>2</sup> | 7.6 | 8.1 | 0.0652 | 0.57 |
| Social roles<sup>3</sup> | 6.7 | 9.1 | 0.0086 | 0.59 |

<sup>1</sup>The minimum and maximum scores for each life habits category are 0 and 10, respectively. A higher score indicates greater independence, less difficulty, and greater satisfaction. The category education of the LIFE-H was not presented in this study, as this category was not applicable for participants.
<sup>2</sup>Day-to-day activities include six life habits categories (communication, mobility, nutrition, physical fitness and psychological well-being, personal care and health, and housing).
<sup>3</sup> Social roles include six life habits categories (responsibilities, interpersonal relationships, community and spiritual life, education, employment, and recreation)
<sup>4</sup>RTE, Relative Treatment Effect.

### 3.2 Measure of Quality of the Environment (MQE)

Table 3 presents the relevant changes between T1 and T2 in environmental factors that facilitated or inhibited the accomplishment of daily habits and social roles according to MQE, as determined by the smallest p-values. Complete results for the MQE are presented in Supplemental Table 2. Briefly, when compared to T1, availability of businesses in the community, such as grocery stores, restaurants and shopping center increased significantly as a facilitator of participants’ social participation (p=0.0013). Electronic communication services (e.g., telephone, internet) were previously identified as a medium facilitator of social participation at T1, but the median score increased towards being a major facilitator (Likert score >2) at T2 (p=0.0626; Table 3). Maintenance services for technical aids were also identified as a factor that had no effect on the accomplishment of daily activities at T1 but increased towards a medium facilitator at T2 (p=0.0187). Finally, rules (e.g., at school, swimming pool, public places, etc.) were identified as a minor obstacle at T1 but increased towards as having no effect on the accomplishment of daily activities at T2 (p=0.0273). All effect sizes were rated small according to Vargha & Delaney (Table 3) (Vargha & Delaney, 2000). The main environmental facilitators and obstacles reported at T2 are presented in Supplemental Figure 3.

**Table 3.**
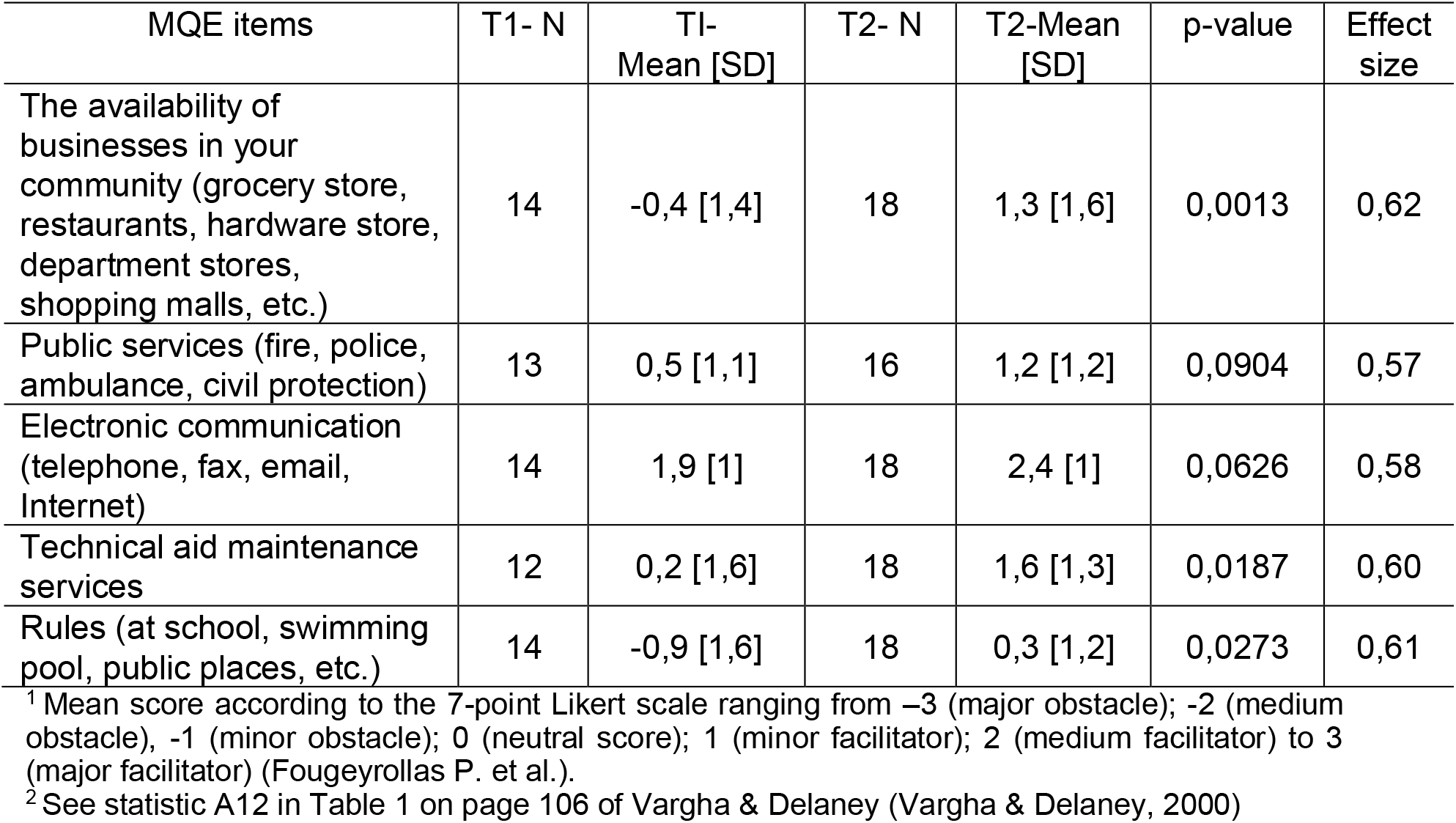
Change of environmental factors that facilitated or inhibited participants’ accomplishment of the day-to-day habits and their social roles according to Measure of Quality of the Environment (MQE)

| MQE items | T1- N | T1- Mean [SD] | T2- N | T2-Mean [SD] | p-value | Effect size |
| --- | --- | --- | --- | --- | --- | --- |
| The availability of businesses in your community (grocery store, restaurants, hardware store, department stores, shopping malls, etc.) | 14 | -0,4 [1,4] | 18 | 1,3 [1,6] | 0,0013 | 0,62 |
| Public services (fire, police, ambulance, civil protection) | 13 | 0,5 [1,1] | 16 | 1,2 [1,2] | 0,0904 | 0,57 |
| Electronic communication (telephone, fax, email, Internet) | 14 | 1,9 [1] | 18 | 2,4 [1] | 0,0626 | 0,58 |
| Technical aid maintenance services | 12 | 0,2 [1,6] | 18 | 1,6 [1,3] | 0,0187 | 0,60 |
| Rules (at school, swimming pool, public places, etc.) | 14 | -0,9 [1,6] | 18 | 0,3 [1,2] | 0,0273 | 0,61 |

<sup>1</sup> Mean score according to the 7-point Likert scale ranging from -3 (major obstacle); -2 (medium obstacle), -1 (minor obstacle); 0 (neutral score); 1 (minor facilitator); 2 (medium facilitator) to 3 (major facilitator) (Fougeyrollas P. et al.).
<sup>2</sup> See statistic A12 in Table 1 on page 106 of Vargha & Delaney (Vargha & Delaney, 2000)

## 4. Discussion

The objective of this study was to describe the change in social participation of adults with SCI after the first two years of the COVID-19 pandemic in Canada. The study highlighted change in facilitators and barriers in the environment of people with SCI that have evolved over the course of the pandemic. Findings also showed increased realization of certain life habits, which together positively influenced the social participation of participants.

### 4.1 Improvement in the accomplishment of certain life habits

On one hand, the achievement of the two main categories of life habits, namely daily activities and social roles, has increased between the first wave of the pandemic and the second wave. Specifically, domains such as mobility, social roles, personal care and health, recreation, and nutrition improved significantly. An improved attainment of life habits was associated with better social participation according to the HDM-DCP. Halvorsen et al. have also reported a strong association between higher levels of participation and quality of life (i.e., life satisfaction and mental health) for people with SCI (Halvorsen et al., 2021). Nevertheless, a study conducted regarding the social participation of people with SCI over the same period as the present study (i.e., December 2020 and June 2021) found divergent results (Robinson-Whelen et al., 2023). Notably, Robinson-Whelen et al. found a decrease in participation in life roles and activities outside of work (Robinson-Whelen et al., 2023). In fact, this study evaluated three life domains outcomes at a single time point. The results may therefore have been compared to the pre-pandemic period, whereas our results compare two measurement times during the pandemic period (Robinson-Whelen et al., 2023). Furthermore, individual characteristics like age, time since injury and social role history may influence the level of social participation (Gross-Hemmi et al., 2021). Future qualitative studies should explore the determinant of social participation and quality of life of people with SCI during and post critical periods like the pandemic context.

### 4.2 Improvement of environmental factors and diminution of barriers to social participation

On the other hand, new environmental factors were identified as facilitators in the MQE questionnaire as the pandemic continued. The opening of businesses in the community, such as grocery stores, restaurants and shopping center, the availability of electronic communication services and maintenance were rated as determinant and significant facilitators of social participation. Previous studies found similar observations, highlighting that the level of perceived environmental barriers is inversely associated with social participation and employment (Tsai et al., 2017). Nevertheless, environmental barriers are still present, which would explain a non-significant improvement in life habits related to associative and spiritual life, physical fitness and physiological well-being, interpersonal relationships. Considering the importance of physical fitness and physiological well-being of people with SCI (Martin Ginis et al., 2012), measures must be put in place to reduce environmental barriers and increase environmental facilitators to enable people with SCI to maintain their social participation.

Several factors may explain the changes in social participation situation as the pandemic evolved. First, people with SCI may have used strategies (Monden et al., 2014) to maintain their social participation during the pandemic. In this regard, a previous study indicated that social support from friends and family was an important coping strategy for people with SCI, especially when faced with events that cause significant changes (Monden et al., 2014). This is consistent with findings in the present study, as support from friends and family was identified as an important environmental facilitator to perform daily activities as the pandemic persisted. Second, the improvement in certain life habits categories, such as mobility, personal care and health, nutrition, and recreation may be explained in part by protective and isolation measures in place at the time of this study. For example, group outdoor sports, cultural and recreational activities that had been previously cancelled, had returned in public places at T2, which may have reduced environmental barriers (Institut national de santé publique du Québec, 2021).

### 4.3 Limitations and strengths

This study includes a small sample of people with SCI using a manual wheelchair. Given that some participants were actively involved in the intervention of the larger study (called AllWheel) during the data collection, they were in contact with a peer (i.e., person with SCI who used a manual wheelchair) who may have influenced social participation during the pandemic (Best et al., 2019). In addition, five participants who did not participate at T1 were added to T2 and one participant who participated in T1 did not participate in T2 (Fortin-Bédard et al., 2022). Furthermore, protective and isolation measures in place during the interview were variable depending on the participant’s area of residence, which may have influenced social participation (Canadian Institute for Health Information, 2022). The psychometric qualities of the LIFE-H version 4.0 have not yet been formally evaluated, but only the version 3.0 (Noreau et al., 2004). In addition, the category education of the LIFE-H was not presented in this study, as this category was not applicable for participants. Finally, the scope of the LIFE-H and MQE questionnaires are inherently limited; e.g., “increased availability of businesses in the community” provides limited information regarding the extent of business availability.

## Conclusion

This study reported changes in the accomplishment of certain life habits and in the environmental factors of individuals with SCI between the first and second waves of the COVID-19 pandemic, influencing the social participation of the Canadian participants in this study. Results suggested that certain life habits categories, such as mobility, personal care and health, nutrition, and recreation have improved during the pandemic, suggesting a better attainment and satisfaction of social participation. Similarly, availability of businesses in the community such as grocery stores, restaurants and shopping center have increased as an environmental facilitator for participants.

## Data Availability

All data produced in the present study are available upon reasonable request to the authors.

## Author Contributions

Conceptualization, F.R and K.L.B; methodology, F.R and K.L.B; formal analysis, J.L; investigation, C.R; writing—original draft preparation, N.F-B, F.N, J.L; writing—review and editing, N.F-B, F.N, J.L, C.R, K.L.B, J.B, S.N.S, K.P.A-N, F.R; supervision, F.R. All authors have read and agreed to the published version of the manuscript.

## Institutional Review Board Statement

The study was conducted in accordance with the Declaration of Helsinki and approved by local Research Ethics Boards.

## Informed Consent Statement

Informed consent was obtained from all subjects involved in the study.

## Data Availability Statement

The data that support the findings of this study are available from the corresponding author, FR, upon reasonable request.

## Acknowledgments

Salary support for Krista Best and François Routhier was provided by the Quebec Health Research Funds (FRQS). Noémie Fortin-Bédard was supported through an Accelerate Mitacs scholarship at the time of the study, and now holds a doctoral training award the FRQS in partnership with the Unité de soutien au système de santé apprenant (SSA) Québec. Shane Sweet is supported by a Canada Research Chair in Participation, Well-Being, and Physical Disability (Tier 2)

