## Supplemental Material for "Change in Social Participation of Adults with Spinal Cord Injury During the First Two Waves of the COVID-19 Pandemic in Canada"

**Supplemental Table 1.** Variables used in the Assessment of Life Habits (LIFE-H) questionnaire to measure each life habits categories [5]

| **Life habits categories[5]** | **Variables[5]** |
|---|---|
| Mobility | - Move short distances (in and out, inside your home)<br>- Moving around outside (street, sidewalk, intersections)<br>- Travel by means of transportation (as a driver or passenger)<br>- Enter and move around the businesses and public and community services in your area (restaurant, grocery store, recreation center, medical clinic)<br>- Get to your main occupation (e.g., work, school, daycare, volunteer work) |
| Associative and spiritual life | - Participate in activities and organizations in your community (social or community groups, religious or spiritual practice) |
| Housing | - Carry out activities related to your residence (e.g., landscaping, maintenance, use of equipment)<br>- Maintain the interior of the residence where you live (e.g., cleaning, laundry, minor repairs) |
| Nutrition | - Prepare your meals<br>- Choose foods for your meals according to your tastes and specific needs (e.g., quantity, freshness, type of food, personal diet, grocery shopping)<br>- Eat your meals (e.g., at home, in a restaurant) |
| Physical fitness and psychological well-being | - Maintain a good physical and mental condition<br>- Engage in physical activities to maintain or improve your physical condition (e.g., walking, individual or group exercises)<br>- Doing activities to ensure your psychological well-being (e.g., yoga, meditation, listening to music) |
| Employment | - Carry out activities related to a job (research, execution of tasks...)<br>- Carry out activities related to an unpaid occupation (e.g., volunteer work, day center, internship) |
| Recreation | - Engage in leisure activities (e.g., art, sports, hobbies, outings, travel) |
| Communication | - Communicate information in different forms (oral, written, physical, electronic)<br>- Access and understand information in different forms (oral, visual, written, electronic) |
| Responsibilities | - Assume your financial, civil and family responsibilities<br>- Shop and use the services in your community<br>- Enforcing your rights (e.g., taking your place, speaking up, expressing your opinion)<br>- Care and support for your family members including your spouse<br>- Ensure the education of your children |
| Personal care and health | - Ensure your personal care (hygiene, appearance, health care)<br>- Dressing and undressing (e.g., choosing and putting on clothes including buttons, zippers, shoelaces, jewelry)<br>- Change your clothes when they are soiled or dirty<br>- Use health services (e.g., medical clinic, hospital, rehabilitation center, dental clinic) |

| | |
|---|---|
| | - Use sanitary facilities (including sinks, toilets and any other equipment needed for disposal) |
| Interpersonal relationships | - Have social, emotional, or intimate relationships with your spouse or family members<br>- Maintain social connections with those around you (e.g., neighbors, colleagues from work, school, or play) |

**Supplemental Figure 1.** Individual scores of the Assessment of Life Habits (LIFE-H) at T2

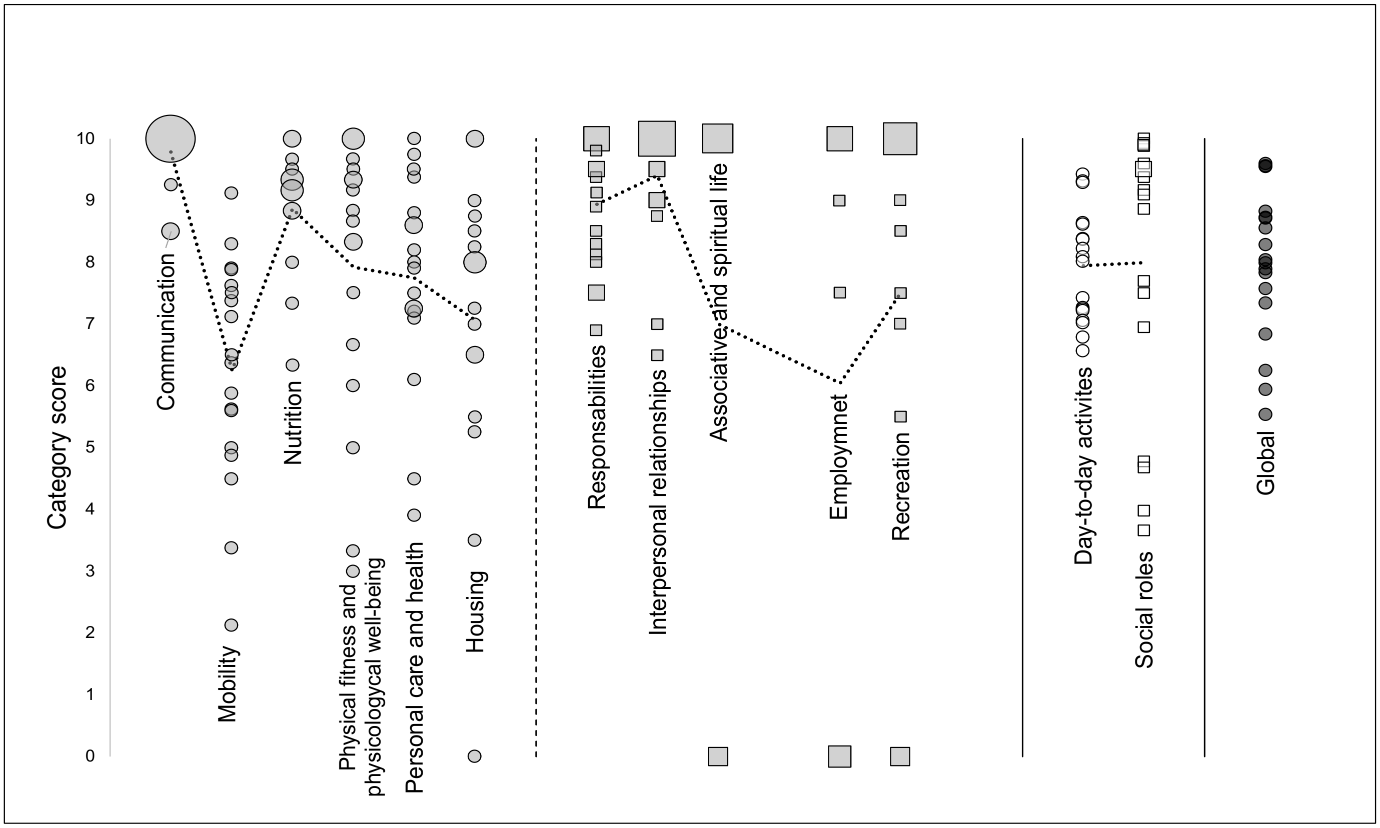


**Figure 1 legend**. Presentation of the 18 participants' social participation according to twelve categories of life habits separated in two categories (day-to-day habits and social roles) in the LIFE-H 4.0 at T2. The life habits categories are presented along the horizontal axis and the scores are presented on the vertical axis. Score can range from 0 to 10 for each item of the questionnaire. Squares were used to illustrate items related to social roles, and circles for items related to day-to-day habits. The size of the shapes is proportional to the quantity of participants having obtained the corresponding score. The lines on the graph indicate the median value of each life habits categories.

**Supplemental Figure 2.** Evolution of the individuals score of Individual scores of the Assessment of Life Habits (LIFE-H) between T1 and T2

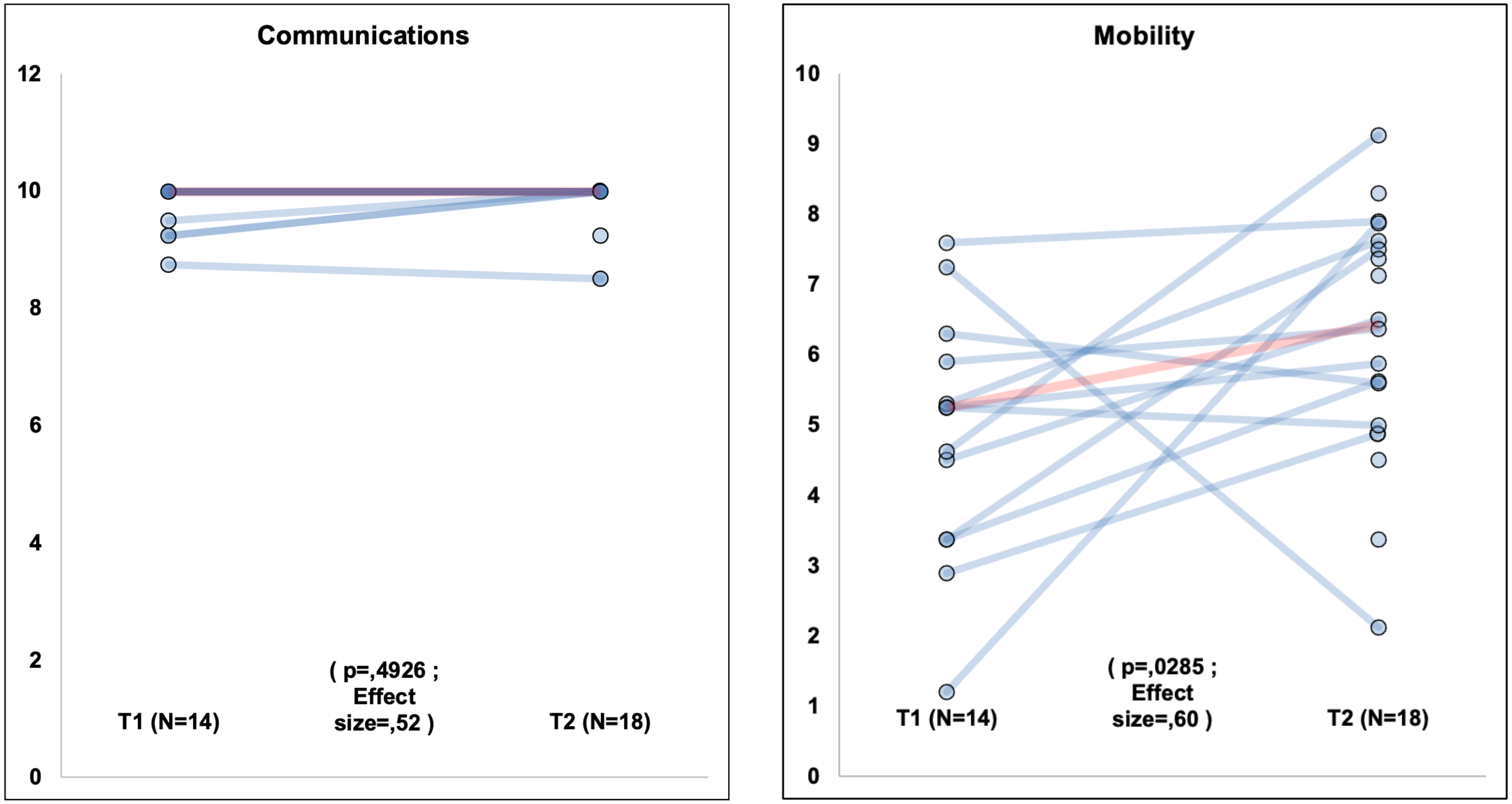

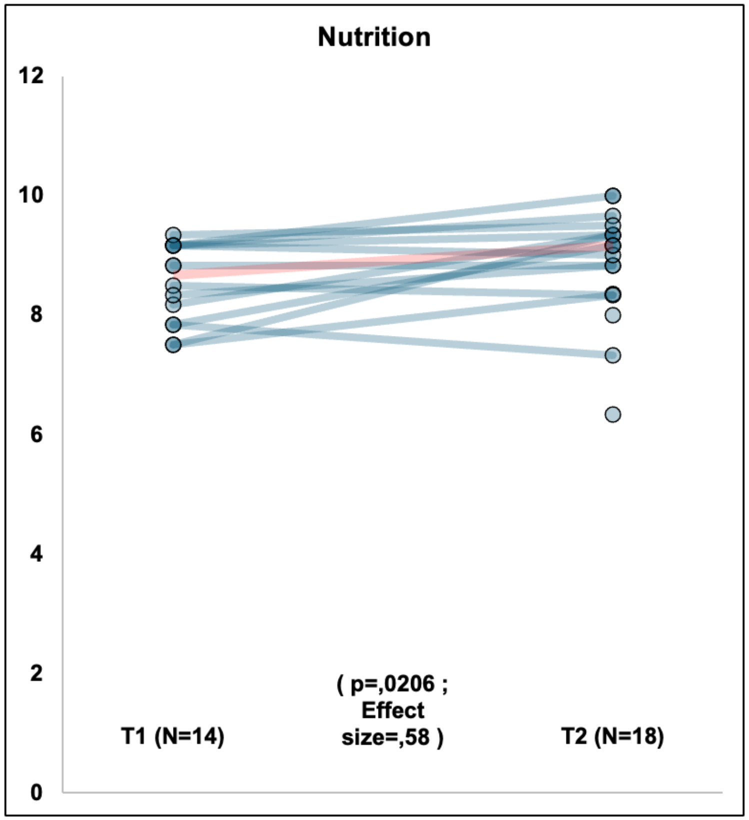
Nutrition
12
10
8
6
4
2
0
( p=,0206 ;
Effect
size=,58 )
T1 (N=14)
T2 (N=18)

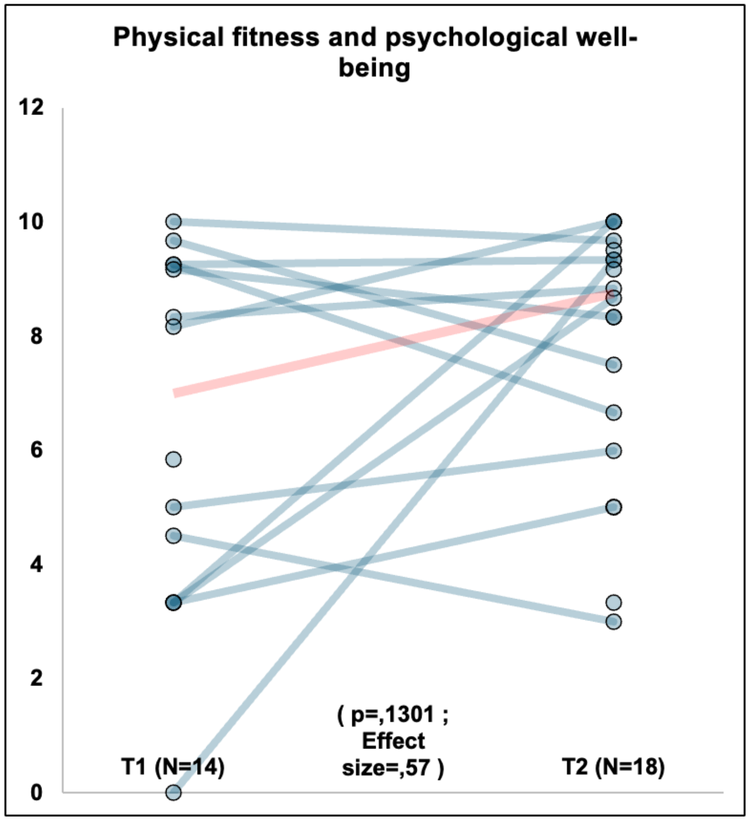
Physical fitness and psychological well-being
12
10
8
6
4
2
0
( p=,1301 ;
Effect
size=,57 )
T1 (N=14)
T2 (N=18)

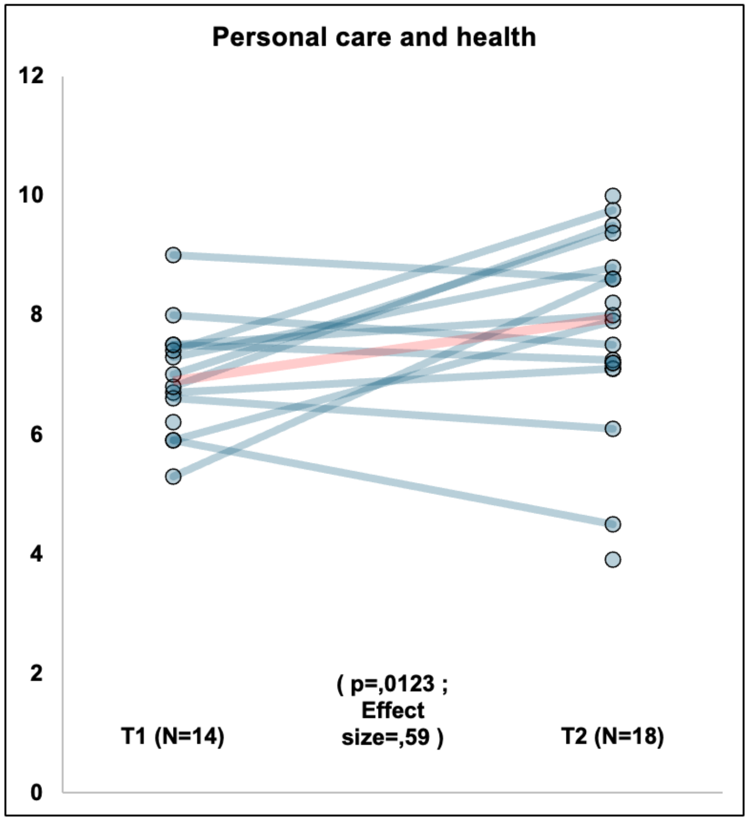
Personal care and health
12
10
8
6
4
2
0
( p=,0123 ;
Effect
size=,59 )
T1 (N=14)
T2 (N=18)

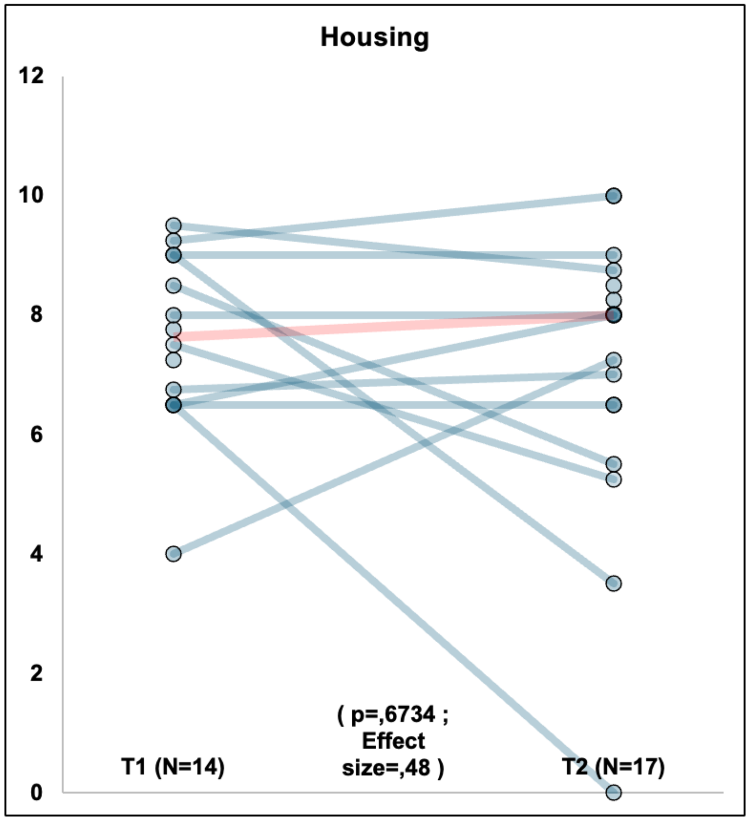
Housing
12
10
8
6
4
2
0
( p=,6734 ;
Effect
size=,48 )
T1 (N=14)
T2 (N=17)

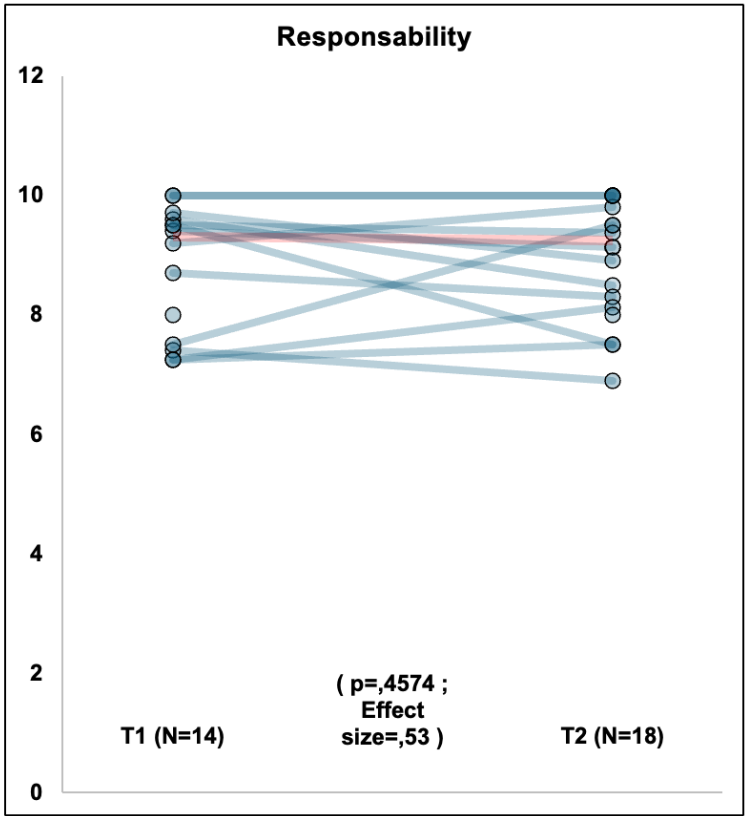
Responsability
12
10
8
6
4
2
0
T1 (N=14)
( p=,4574 ; Effect size=,53 )
T2 (N=18)

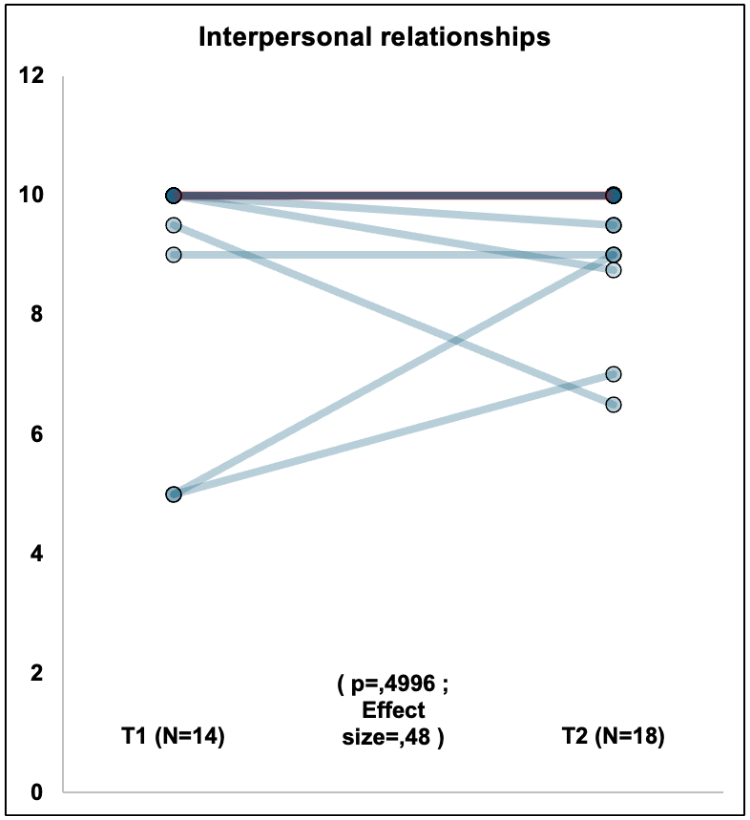
Interpersonal relationships
12
10
8
6
4
2
0
T1 (N=14)
( p=,4996 ; Effect size=,48 )
T2 (N=18)

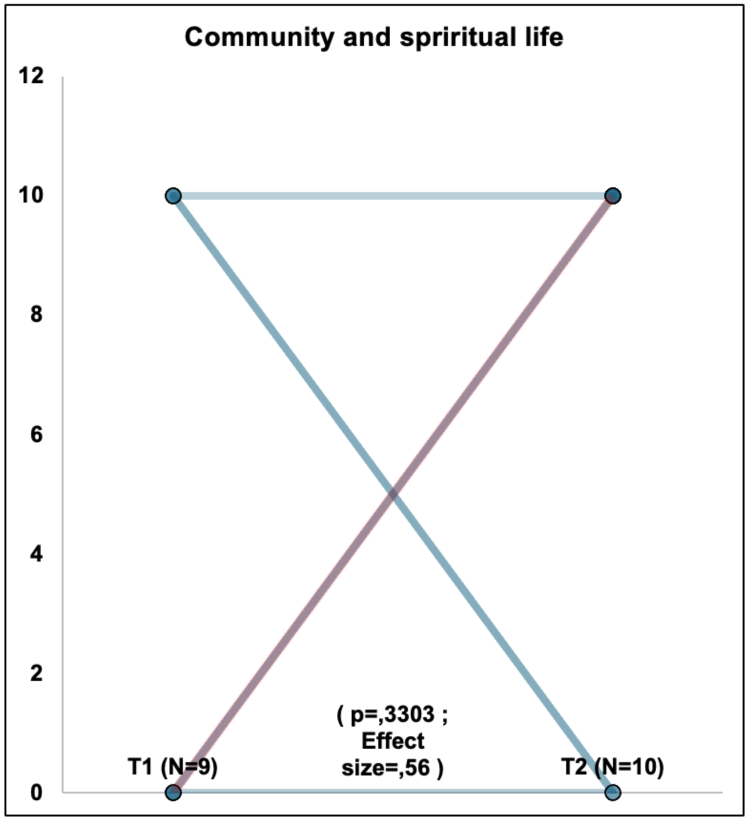
Community and spriritual life
12
10
8
6
4
2
0
T1 (N=9)
( p=,3303 ;
Effect
size=,56 )
T2 (N=10)
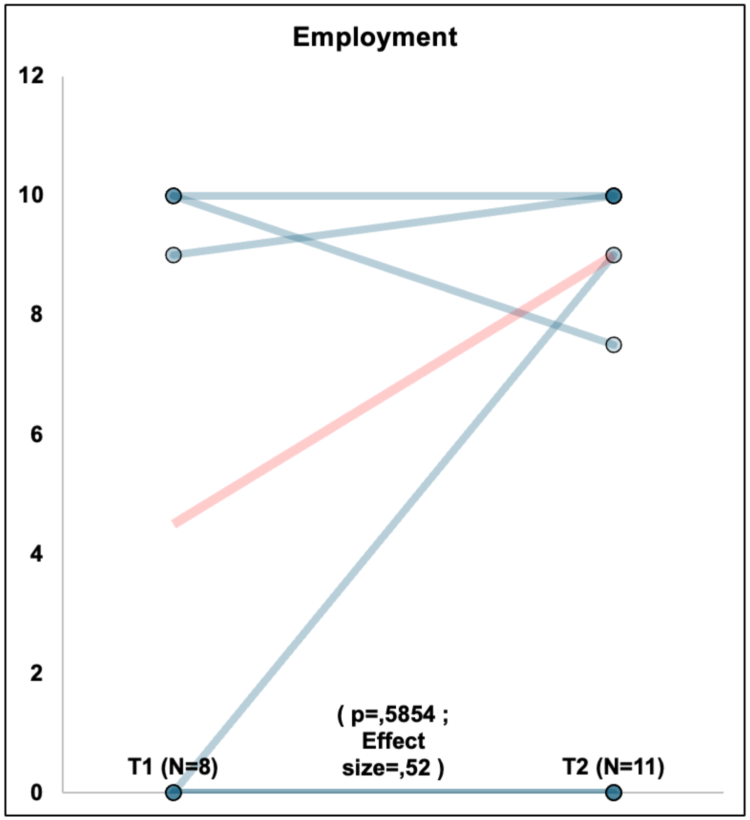
Employment
12
10
8
6
4
2
0
T1 (N=8)
( p=,5854 ;
Effect
size=,52 )
T2 (N=11)

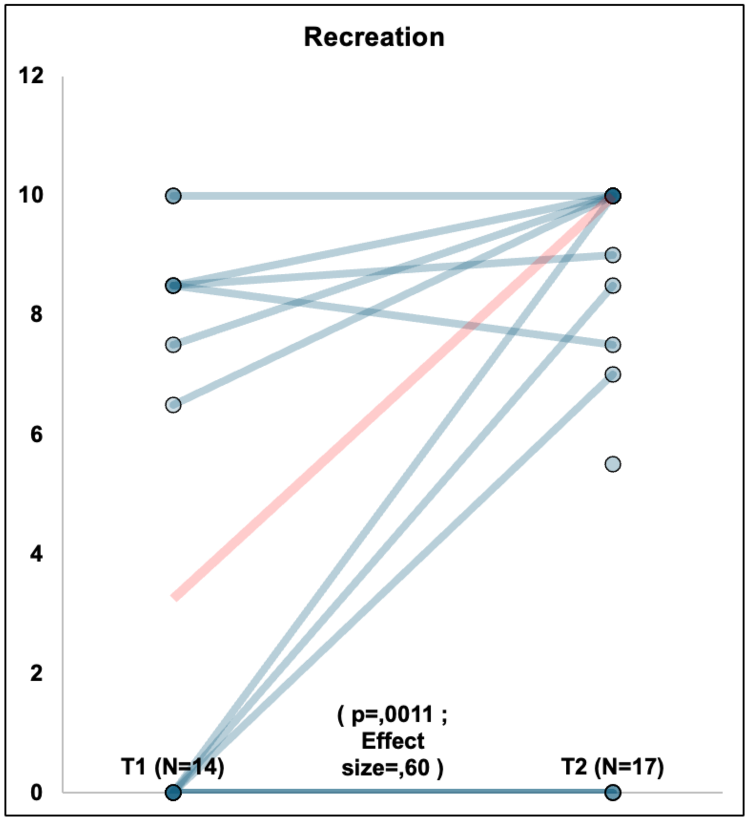
Recreation
12
10
8
6
4
2
0
T1 (N=14)
( p=,0011 ;
Effect
size=,60 )
T2 (N=17)

**Supplemental Table 2.** Evolution of the environmental factors that facilitated or inhibited participants' accomplishment of the day-to-day habits and their social roles according to Measure of Quality of the Environment (MQE)

| MQE items | T1- N | TI-Mean [SD] | T2- N | T2-Mean [SD] | P-value | Effect size | Common language effect size statistic (CL) |
|---|---|---|---|---|---|---|---|
| Your family situation (living alone, with a spouse, or with children) | 14 | 1,7 [1,6] | 18 | 1,8 [1,5] | 0,9366 | 0,50 | |
| Support from members of your family or close friends who take the place of family (presence, physical assistance, household assistance, encouragement) | 14 | 1,4 [1,2] | 18 | 1,9 [1,2] | 0,1915 | 0,56 | |
| Support from your friends | 13 | 1,2 [1,2] | 18 | 1,4 [1,1] | 0,7540 | 0,52 | |
| Support from your neighbours | 14 | 0,4 [0,9] | 17 | 0,5 [0,9] | 0,8614 | 0,51 | |
| Support from your colleagues at work, school, or place of principal occupation | 4 | 1 [1,8] | 9 | 1 [1,1] | 0,9455 | 0,50 | |
| The attitudes of your family or close friends who take the place of family toward you | 14 | 1 [1,5] | 17 | 1,3 [1,4] | 0,5028 | 0,52 | |
| The attitudes of your friends toward you | 13 | 1,2 [1,2] | 18 | 1,4 [1,1] | 0,7326 | 0,51 | |
| The attitudes of your neighbours toward you | 14 | 0,4 [0,9] | 17 | 0,7 [1] | 0,5899 | 0,52 | |
| The attitudes of your service providers (public services agents, salespeople, cashiers, etc.) toward you | 12 | 1,1 [1,4] | 17 | 0,9 [1,5] | 0,7413 | 0,49 | |
| Availability of jobs in your community | 6 | -0,5 [1,4] | 10 | -0,4 [1,2] | 0,7893 | 0,51 | |
| The characteristics of your work environment and your working conditions | 2 | 1,5 [2,1] | 5 | 1 [2] | 0,6996 | 0,47 | |
| Your personal income | 14 | 0,5 [1,9] | 18 | 0,8 [2,1] | 0,5097 | 0,52 | |
| Financial compensation programs (subsidized rent, disabiliy compensation, direct payments, etc.) | 12 | 1,4 [1,2] | 16 | 1 [1,7] | 0,3649 | 0,47 | |
| Socio-economic services (fiscal programs, family allocations, unemployment insurance) | 10 | 1,1 [1,4] | 14 | 0,9 [1] | 0,7340 | 0,49 | |
| The availability of businesses in your commnity (grocery store, restaurants, hardware store, department stores, shopping malls, etc.) | 14 | -0,4 [1,4] | 18 | 1,3 [1,6] | 0,0013 | 0,62 | Small |
| The services offered by businesses in your community | 14 | 1,4 [1,2] | 18 | 1,3 [1,5] | 0,9500 | 0,50 | |

| | | | | | | | |
|---|---|---|---|---|---|---|---|
| External attendant services other than those provided by your family and close friends (escort, interpreter, etc.) | 3 | 1 [1,7] | 4 | -0,8 [1,5] | 0,1615 | 0,39 | Small |
| Home care services other than those provided by your family and close friends | 7 | 1,4 [1,5] | 13 | 1,8 [1,9] | 0,4556 | 0,54 | |
| The health services in your community (hospital, medical clinic, dentist, etc.) | 13 | 0,7 [1,5] | 18 | 1,3 [1,6] | 0,2423 | 0,55 | |
| Social integration support services (social work, residential resources, etc.) | 1 | 0 [0] | 7 | 0,7 [1,6] | | | |
| Day care services (including home day care) and in-school child care | 3 | 0,7 [1,2] | 4 | 0,5 [1] | 0,8694 | 0,48 | |
| Other child care services (providing time to rest and helping out) | 1 | -2 [-2] | 5 | 1,6 [1,5] | | | |
| Educational services | 4 | -0,5 [2,1] | 5 | 0,4 [0,5] | 0,4419 | 0,58 | Small |
| The personal vehicle that you use | 14 | 2,4 [0,9] | 16 | 2,2 [1,1] | 0,5747 | 0,48 | |
| Public transportation services in your community (schedule, stops, frequency, route, etc.) | 7 | -0,1 [0,4] | 10 | -0,1 [0,6] | 0,9154 | 0,51 | |
| Adapted transportation services (schedule, stops, frequency, route, etc.) | 10 | 0,8 [1,3] | 14 | 0,3 [1,4] | 0,4160 | 0,46 | |
| Long distance transportation services (train, bus, plane) | 11 | -0,5 [1,3] | 15 | 0,1 [0,3] | 0,2500 | 0,55 | |
| Radio and television (access, quality of information, subtitling, sign language, etc.) | 14 | 1,4 [1,2] | 17 | 1,2 [1,4] | 0,9287 | 0,50 | |
| Television media services | 14 | 1,1 [1,1] | 18 | 1,5 [1,3] | 0,3758 | 0,54 | |
| Public services (fire, police, ambulance, civil protection) | 13 | 0,5 [1,1] | 16 | 1,2 [1,2] | 0,0904 | 0,57 | Small |
| Electronic communication (telephone, fax, E-mail, Internet) | 14 | 1,9 [1] | 18 | 2,4 [1] | 0,0626 | 0,58 | Small |
| Municipal services (road maintenance, snow removal, garbage pick-up, etc.) | 14 | 0,8 [1,2] | 18 | 1,3 [1,3] | 0,1860 | 0,56 | |
| Community and cultural services in your community (cultural, sports, religious organizations) | 11 | 0,8 [1,6] | 18 | 0,2 [0,9] | 0,2583 | 0,45 | |
| Availability of technical aids (wheelchairs, orthosis, writing assistance, guide-dogs, etc.) | 14 | 1,6 [1,9] | 18 | 1,8 [1,6] | 0,8401 | 0,51 | |
| Use of technical aids | 14 | 1,9 [1,8] | 18 | 2,1 [1,3] | 0,8846 | 0,51 | |
| Technical aid maintenance services | 12 | 0,2 [1,6] | 18 | 1,6 [1,3] | 0,0187 | 0,60 | Small |
| Means of participating in decision-making in your community | 13 | 0,6 [1,4] | 18 | 0,2 [0,6] | 0,3124 | 0,45 | |
| Actions of advocacy organisations | 13 | 0,7 [1] | 17 | 0,4 [0,8] | 0,4388 | 0,47 | |
| Governmental and administrative procedures | 13 | 1 [1,4] | 17 | 1,1 [1,3] | 0,9838 | 0,50 | |

| | | | | | | | |
|---|---|---|---|---|---|---|---|
| Equal opportunity programs (access to education, labour market, etc.) | 9 | -0,1 [0,8] | 15 | 0,5 [0,9] | 0,1751 | 0,55 | |
| Responsibilities and coherence of diverse government levels | 13 | 0,2 [0,7] | 18 | 0,4 [1,1] | 0,7547 | 0,51 | |
| Administrative procedures (bureaucracy, forms, etc.) | 13 | -0,6 [1,3] | 17 | -0,6 [1,3] | 0,9405 | 0,50 | |
| Rules (at school, swimming pool, public places, etc.) | 14 | -0,9 [1,6] | 18 | 0,3 [1,2] | 0,0273 | 0,61 | Small |
| Conventions (eligibility criteria, collective agreement, etc.) | 11 | 0 [0,4] | 16 | 0,1 [0,4] | 0,7205 | 0,51 | |
| Law enforcement (smoking laws, parking laws, etc.) | 13 | 0,2 [0,7] | 18 | 0,6 [1,1] | 0,2495 | 0,54 | |

**Supplemental Figure 3**. Main Facilitators and Barriers Identified in the Measure of Quality of the Environment (MQE) during T2.

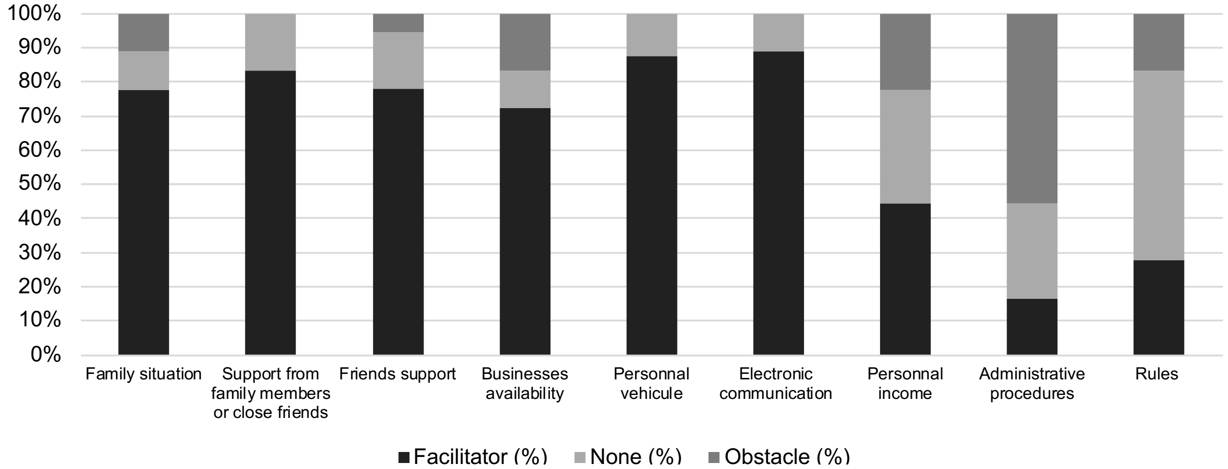


**Figure 3 legend.** Presentation of the main environmental factors that facilitated or inhibited participants' accomplishment of the day-to-day habits and their social roles, and consequently their social participant during T2. The percentage is the proportion of participants with the corresponding score.
